# Phase 1 Trial of Cyclosporine for Hospitalized Patients with COVID-19

**DOI:** 10.1101/2021.06.10.21258714

**Authors:** Emily A. Blumberg, Pablo Tebas, Ian Frank, Amy Marshall, Anne Chew, Elizabeth A. Veloso, Alison Carulli, Walter Rogal, Avery L. Gaymon, Aliza H. Schmidt, Tiffany Barnette, Renee Jurek, Hooman Noorchashm, Wei-Ting Hwang, Julia Han Noll, Joseph A. Fraietta, Carl H. June, Elizabeth O. Hexner

## Abstract

Coronavirus Disease 2019 (COVID-19) remains a global health emergency with limited treatment options, lagging vaccine rates and inadequate healthcare resources in the face of an ongoing calamity. The disease is characterized by immune dysregulation and cytokine storm. Cyclosporine A (CSA) is a calcineurin inhibitor that modulates cytokine production and may have direct antiviral properties against coronaviruses. To test whether a short course of treatment was safe in COVID-19 patients, we treated 10 hospitalized, oxygen requiring, non-critically ill patients with CSA at a starting dose of 9mg/kg/day. Five patients experienced adverse events, none were serious, and transaminitis was most common. No subject enrolled in this trial required intensive care unit (ICU)-level care and all patients were discharged alive from the hospital. Further, CSA treatment was associated with significant reductions in serum cytokines and chemokines important in COVID-19 hyper-inflammation, including CXCL10. In conclusion, short courses of CSA appear safe and feasible in COVID-19 patients requiring oxygen and therefore, may be a useful adjunct in resource-poor or resource-limited health care settings.

## Introduction

SARS-CoV-2, the causative agent of COVID-19, is a novel coronavirus that induces an acute respiratory disease with systemic complications that range from minimally symptomatic, self-limited disease to critical illness and death. The COVID-19 pandemic is an ongoing global health crisis. Although vaccine development has been rapid, safe and effective, treatment of disease has largely suffered from a paucity of effective antiviral drugs and variable impact of anti-inflammatory agents, some of which are both costly and associated with long-term immunosuppression. Global infection rates continue to rise, vaccines are lagging, and case fatality rates vary globally but remain in the 1-10% range. As of June 2021, global infections exceeded 170 million, with over 3.7 million dead [1]. Severe pneumonia occurs in approximately 15% of cases and drives mortality. Access to critical care resources such as ventilators or even supplemental oxygen remains an unmet need in some areas more than a year into the pandemic.

COVID-19 is characterized by immune dysregulation or cytokine storm, an orchestrated response involving infected cells, effector T cells, macrophages and other innate immune cells, and the cytokines/chemokines produced that collectively result in widespread lung inflammation [2]. Hospitalized patients with severe COVID-19 exhibit high serum levels of interleukin (IL)-2, IL-7, IL-10, granulocyte colony-stimulating factor (G-CSF), tumor necrosis factor (TNF), C-X-C motif chemokine ligand 10 (CXCL10), monocyte chemoattractant protein 1 (MCP1), and macrophage inflammatory protein (MIP)1 α [3][4]. Systemic elevations of these cytokines, C-reactive protein (CRP), and ferritin accompanied by lymphopenia are frequently observed in patients with COVID-19, and are also hallmarks of patients with hemophagocytic lymphohistiocytosis), also referred to as macrophage activation syndrome (HLH) [5].

Calcineurin inhibitors are a class of non-cytotoxic immunosuppressive drugs that selectively impair T cell function by blocking Nuclear factor of activated T cells (NFAT) signaling and downstream cytokine production. A drug screen conducted at the University of Pennsylvania identified cyclosporine as an active antiviral agent in human lung cells, blocking entry of SARS-CoV-2 [6]. Currently cyclosporine A (CSA), tacrolimus and sirolimus are widely used to prevent rejection in solid organ transplant and for the treatment of arthritis and psoriasis; CSA is also the backbone of most protocols treating HLH. Given the shared inflammatory pathways seen in both HLH and the immune dysregulation seen in severe COVID-19 infection, we hypothesized that CSA could be an effective anti-inflammatory agent for the treatment of COVID-19-associated immunopathology. Furthermore, CSA is not myelosuppressive and has also been shown to have direct antiviral effects by inhibiting coronavirus replication.[6, 7]. Thus, we asked whether, if properly timed in patients with COVID-19, CSA would be sufficiently safe so that ultimately it could serve as a broad-spectrum inhibitor to help control SARS-CoV-2 infection, decrease severity of cytokine storms and improve outcomes. This early intervention with a well characterized and approved medication might be of particular importance in resource-poor or resource-limited areas.

## Methods

### Design and Study Population

This was a Phase 1, single-site, single-arm, open label design of a short course CSA treatment of hospitalized patients with COVID-19. Eligible patients were adults over the age of 18, admitted to hospital with laboratory confirmation of SARS-CoV-2 infection, requiring supplemental oxygen, with an estimated creatinine clearance >50 mL/minute. Patients were excluded if admitted to the ICU at time of enrollment, had an additional, active unconrolled infection with a non-COVID-19 pathogen, had an active malignancy, or were on chronic immune suppressive medications for other indications. They were also excluded if they had received prior treatment with immunomodulators or immunosuppressant drugs within 5 half-lives or 30 days of consent, such as IL-1, IL-6 or TNF inhibitors, or Janus kinase (JAK) inhibitors. Also excluded were pregnant or lactating women and patients receiving investigational vaccines for SARS-CoV-2. Dexamethasone as standard of care therapy for SARS-CoV-2 and use of inhaled steroids were allowable.

This investigator-initiated trial protocol was approved by the Institutional Review Board (IRB) at the University of Pennsylvania and was overseen by the Center for Cellular Immunotherapies (IND#149997, ClinicalTrials.gov identifier: NCT04412785). Informed consent was obtained from each patient.

### Intervention

The study treatment, cyclosporine (modified, Gengraf^®^) capsules (25 and 100mg) was given at an initial starting dose of 9mg/ kg/day orally divided into dosing every 12 hours, with a maximum dose of 400mg/dose for all subjects. An oral solution (100mg/mL) or intravenous (IV) formulation (Sandimmune^®^; 3mg/kg/day given by continuous infusion) was also available for patients unable to swallow capsules. Therapeutic drug monitoring was performed on day 2 and every Monday, Wednesday and Friday during active dosing. Subsequent cyclosporine dosing was adjusted to target a trough level of 200 to 300ng/mL without a maximum dose level. The intended duration of administration was up to 14 days with planned treatment discontinuation if mechanical ventilation was required. Treatment was held for significant elevations in creatinine or transaminases and discontinued for all patients at the time of hospital discharge.

### Outcomes

This was a study to assess the safety of CSA in subjects with COVID-19, as measured by treatment-related adverse events (AEs), on study ICU transfer, secondary infections, and need for mechanical ventilation or increase in supplemental oxygen requirements. Samples for exploratory measurements (e.g., serum cytokine levels) were also collected.

This safety study also defined the following events as ones that would trigger a pause of the study: moderate to severe superinfection, severe microangiopathy or Posterior Reversible leuko-Encephalopathy syndrome (PRES), also known as Reversible Posterior Leukoencephalopathy syndrome (RPLS), and subject death.

### Laboratory and Correlative Analyses

Clinical laboratory tests at screening/enrollment and post-CSA administration included complete blood counts and assessment of C-reactive protein (CRP) levels. Routine blood analysis was performed using a fully automated cell counter in the University of Pennsylvania Hematology Laboratory. CSA concentration was measured in whole blood samples using a validated Liquid Chromatography-Tandem Mass Spectrometry method.

For cytokine measurements, serum samples stored at −80°C were thawed and centrifuged at 2,000 × g for 5 minutes. The LEGENDplex™ COVID-19 Cytokine Storm Panels 1 and 2 (BioLegend) were used to determine cytokine concentrations, with sample analysis performed using an LSRFortessa flow cytometer (BD Biosciences). Serum was diluted 1:2 and the procedure was carried out according to the manufacturer’s instructions with the addition that capture beads were inactivated in 4% paraformaldehyde for 15 minutes at room temperature and washed once in wash buffer after completion of the staining protocol in a Class II Biosafety Cabinet under BSL-2+ conditions.

### Sample Size and Statistical Analysis

The initial trial was designed to enroll approximately 25 patients for a target of maximum 20 patients treated with CSA. We estimated that 20% of enrolled subjects would not receive CSA, primarily due to unanticipated rapid clinical deterioration or improvement. The sample size would enable us to provide a reasonable precision to rates of adverse events: a sample size of 20, the half-width of 90% exact confidence interval for an AE rate would be no more than 20%. After study activation, accrual rate tracked with infection rates, and when infection rates were declining, lengths of hospital stays were decreasing and vaccines became widely available a decision was made to close the study. With a reduced sample size of 10 patients, we have probability of 0.9 that the true AE rate would be <26%, <40%, if 0 or 1 AEs are observed. Descriptive statistics were computed for all endpoints.

Serum cytokine levels were analyzed between time points using two-sided paired Student’s *t*-tests. Statistical analyses were performed using Prism 8 GraphPad (GraphPad Software). *P* values < 0.05 were considered to be statistically significant.

## Results

### Patients

Eleven patients consented to the study and 10 patients were treated (**Figure 1**). The median age was 57.5 years (**Table 1**). All were oxygen requiring with a median National Early Warning Score of 3 (range 2-8).

**Table 1.** Demographic and Clinical Characteristics of the Patients at Baseline.

|  |  |
| --- | --- |
| Number of Patients | *n* = 10 |
| Median Age - years (range) | 57.5 (36 - 63) |
| Sex |  |
| Females - number (%) | 5 (50%) |
| Males - number (%) | 5 (50%) |
| Race - number (%) |  |
| American Indian | 1 (10%) |
| Black or African American | 3 (30%) |
| White | 5 (50%) |
| Ethnic Group - number (%) |  |
| Hispanic or Latino | 2 (20%) |
| Median Days from Symptom Onset - number (range) | 1 (1 - 3) |
| One or More Relevant Comorbidities - number (%)* | 7 (70%) |
| National Early Warning Score - median (range) | 3 (2 - 8) |

\*Obesity (6), Diabetes (5), hypertension (4), asthma/obstructive lung disease (2), cancer history (1), cardiomyopathy (1)

**Figure 1:**
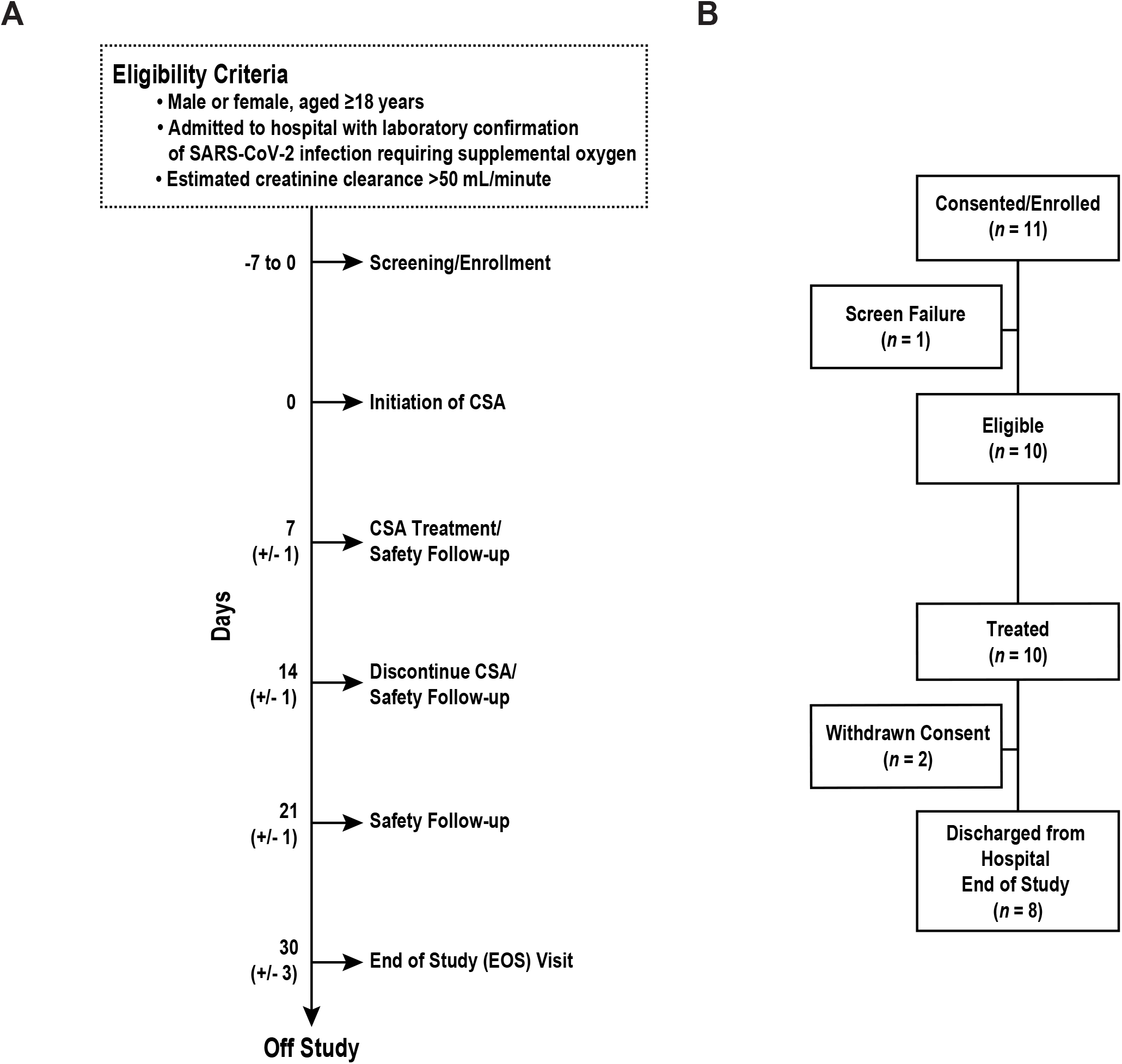
Protocol design and consort diagram for trial of CSA in hospitalized patients with COVID-19. (**A**) Eligibility criteria and protocol schema for screening, CSA treatment and follow-up safety assessments. If hospital discharge occurred prior to day 14, treatment was discontinued at discharge. (**B**) CONSORT diagram indicating the number of patients screened and enrolled in the study.

### Cyclosporine Levels

While the optimal dose is unknown for this indication, for this initial safety study a standard transplant target dose was selected to achieve a trough level of 200 to 300ng/ml. All patients received only the oral capsule formulation of CSA. The median number of days of treatment was 4 (range 2 –6), median doses received was 8 (range 3 –11). Trough levels (**Figure 2**) ranged from 83 to >500 ng/ml. The target trough level was achieved in 80% of patients, and 80% of patients had dose modifications based on therapeutic drug monitoring.

**Figure 2:**
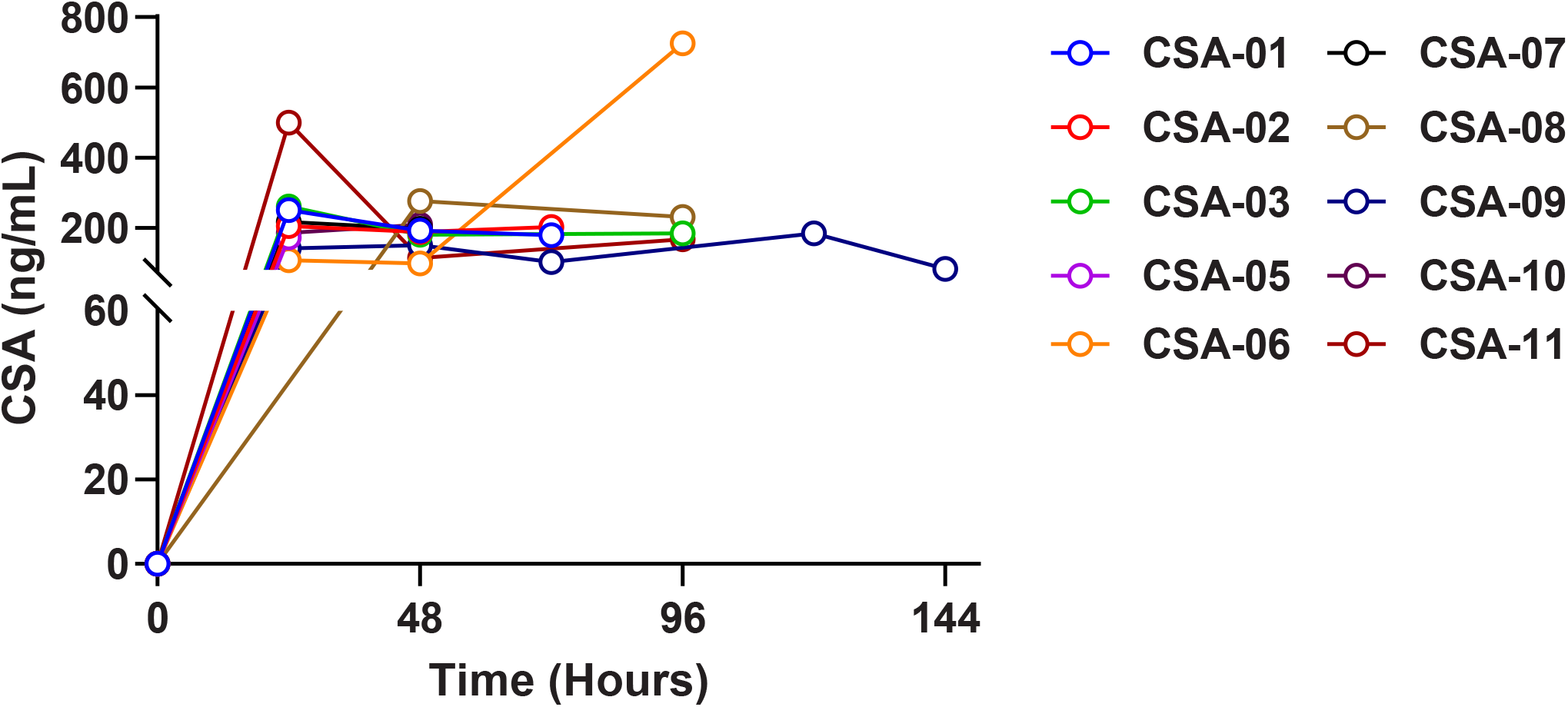
Serial trough levels of CSA following intervention. CSA was administered at a starting dose of 9mg/kg/day and adjusted to target a trough level of 200 to 300ng/mL. Each differentially colored line represents CSA trough levels over time for an individual subject.

### Concomitant Treatments

The trial opened on June 22, 2020, when remdesivir was still being investigated and continued through its approval as well as the introduction of dexamethasone as part of our standard institutional treatment protocol. All patients received both remdesivir and dexamethasone as part of standard of care treatment.

### Safety

Five patients (50%, CI, 22% to 78%) experienced AEs, **(Table 2**). Transaminitis was the most common AE, there was one event each of headache and creatinine increase. Two patients discontinued treatment due to AEs. No patients required ICU-level care and all patients were discharged alive from the hospital. There were no events of PRES/RPLS or microangiopathy, and no serious AEs.

**Table 2.** Adverse Events.

| *Grade* | *1* | *2* | *3* | *Total* |
| --- | --- | --- | --- | --- |
| Alanine aminotransferase increased | 1 | 0 | 1 | 2 |
| Aspartate aminotransferase increased | 0 | 0 | 1 | 1 |
| Creatinine increased | 0 | 1 | 0 | 1 |
| Hypertriglyceridemia | 1 | 0 | 0 | 1 |
| Hypoalbuminemia | 1 | 0 | 0 | 1 |
| Hypomagnesemia | 1 | 0 | 0 | 1 |
| Headache | 0 | 1 | 1 | 2 |
| **Total** | **4** | **2** | **3** | **9** |

### Reduction in Pathogenic Inflammation

Serum specimens were analyzed to quantify circulating pro-inflammatory cytokine/chemokine s levels using multiplex bead-based immunoassays. In most of the 10 patients accrued (*n* = 6), samples were collected twice, typically from day −7 to 0 (baseline) and at day 3 post-CSA administration. A subset of patients (*n =* 5) had cytokine measurements performed more than once after enrollment. At baseline, we detected high levels of multiple pro-inflammatory cytokines and chemokines known to characterize COVID-19-associated hyper-inflammation [3] (**Figure 3A**). Significant or near significant reductions in CXCL10, IL-10, IL-7 and IL-8 were observed on day 3 following CSA administration, relative to pre-treatment time points (**Figure 3B**). The overall inflammatory cytokine signature continued to decrease from day 3 to day 7 post-CSA administration (**Figure 3A**), in association with reductions in body temperature (**Figure 3C**) and non-cardiac CRP levels (**Figure 3D**). Reductions in pro-inflammatory cytokine production coincided with increased white blood cell (WBC; **Figure 3E**) and absolute lymphocyte counts (ALC; **Figure 3F**).

**Figure 3:**
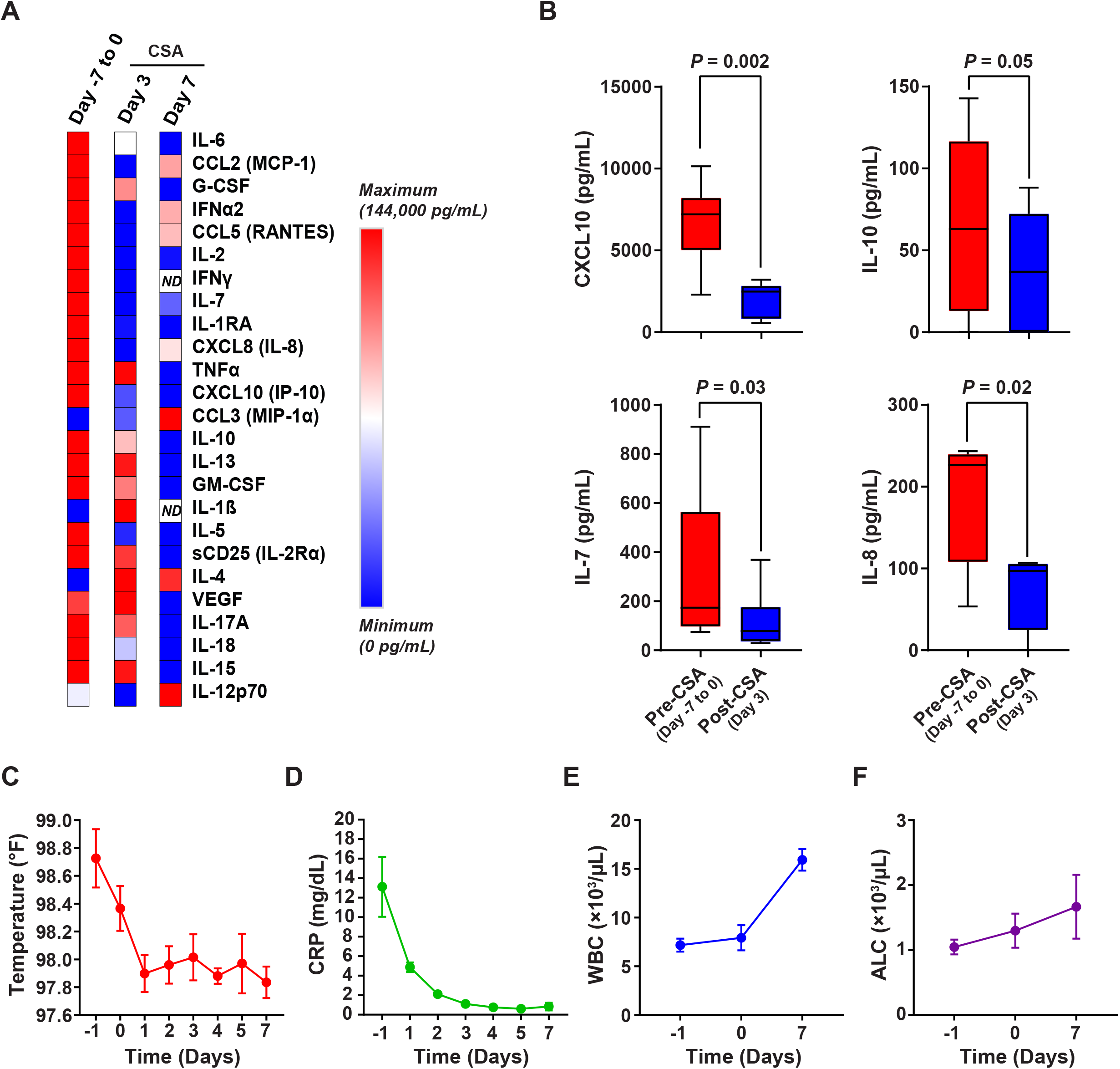
Characterization of the hyper-inflammatory state in hospitalized COVID-19 patients treated with CSA. (**A**) Heat map showing changes in expression of several serum inflammatory cytokines/chemokines over time in hospitalized subjects with COVID-19. *ND* = not detected. (**B**) Box plots depicting baseline and post-CSA treatment (day 3) serum levels of select pro-inflammatory mediators implicated in the COVID-19 cytokine storm in evaluable patients. The boxes depict the first and third quartiles and band within boxes indicates the median. Maximum and minimum data points are depicted by whiskers. *P* values were calculated using a parametric two-sided Student’s *t* test for paired samples. Clinical assessments such as (**C**) body temperature (**D**) inflammation status (CRP levels) (**E**) white blood cell (WBC) counts and (**F**) absolute lymphocyte counts (ALC) are shown across all longitudinal time points for *n* = 10 patients with COVID-19 treated with CSA. Error bars depict the SEM.

## Discussion

CSA is approved by the US FDA for prophylaxis of organ rejection in solid organ transplant recipients, treatment of rheumatoid arthritis and severe psoriasis. CSA, given for a short course to hospitalized patients with COVID-19 and requiring oxygen, is feasible and appears safe. AEs were consistent with the known safety profile of CSA in other populations [8][9, 10]. The majority of AEs were mild and no stopping rules were met. Notably, all patients improved and none required mechanical ventilation. This initial study was not designed to determine efficacy as to whether CSA impacted outcomes. An ideal point of intervention for CSA may be after a sufficient amount of time has passed following acquisition that the humoral immune response to SARS-CoV-2 has been primed, but potentially aberrant immune activation has not yet occurred. This is in contrast to immunologically naïve patients, because administration of CSA prior to infection can be deleterious [15][10].

One advantage to the use of CSA is the cost effectiveness of this intervention for resource limited settings. Drug acquisition costs are low and this medication is widely available in both pill and liquid formulations, making it easy to administer orally to diverse populations. It is likely that lower doses can be given without a need for therapeutic drug monitoring. Because remdesivir is a substrate for cytochrome P450 3A4 (CYO3A4), organic anion transporting polypeptide 1B1 (OATP1B1), and P-glycoprotein 1 (P-gp) *in vitro* (reviewed in [11, 12]), there is a potential for a significant drug interaction with CSA. However, based on our experience, this interaction appears to be manageable. Additionally, given the short recommended duration of therapy with remdesivir and the likelihood that short courses of cyclosporine will also be beneficial, we do not anticipate significant AEs related to the co-administration of these medications. More experience with diverse populations will help further clarify this issue.

Several reports have revealed that the hyper-inflammatory response associated with COVID-19 is a major cause of disease severity and death. In this study, we detected decreased levels of several pro-inflammatory cytokines/chemokines, including CXCL10, following CSA administration compared to baseline time points. Continuously high levels of CXCL10 have been previously associated with increased viral load, loss of respiratory function, lung injury and a fatal outcome in SARS-CoV-2 infection [13]. Furthermore, it has been proposed that CXCL10 may mediate the aberrant immune response that drives the duration of mechanical ventilation in COVID-19 patients with acute respiratory distress syndrome (ARDS)[14]. CXCL10 has also been associated with disease severity of H5N1, H1N1, SARS-CoV, and MERS-CoV [15-18]. Notably, CSA is a robust inhibitor of CXCL10-induced NFATc1 activation and this is a mechanism by which CXCL10 regulates the recruitment of inflammatory cells in rheumatoid arthritis [19]. Thus, modulation of CXCL10 through short course CSA treatment may be a promising therapeutic approach to prevent progression to COVID-19 related ARDS.

There are several limitations to this study. Given the variable epidemiology of COVID-19 and the changing therapeutics during our study period, our enrollment was impacted and the number studied is small. The small numbers also affected our ability to estimate whether there was a significant clinical effect for our patients, independently of those related to corticosteroids and remdesivir.

In summary, we show that CSA is a safe and potentially effective therapeutic intervention for patients with SARS-CoV-2 infection. Its anti-inflammatory properties, wide availability, safety and low cost make it a particularly attractive modality for use in resource-limited settings. A prospective randomized controlled trial testing the efficacy of cyclosporine for the treatment of COVID-19 pneumonia is registered in Spain [20]. Based on our results further large-scale trials are warranted to explore the safety and benefits of this intervention globally.

## Data Availability

Any data that support the findings of this study are available from the corresponding author upon reasonable request.

## Acknowledgements

JAF and CHJ receive research funding supported in part by a grant from the Parker Institute for Cancer Immunotherapy. The authors would like to thank Drs. James Riley and E. John Wherry who established and operationalized the COVID-19 Processing Unit (CPU) that was launched in March of 2020 to facilitate SARS-CoV-2 and COVID-19 research at the University of Pennsylvania. Sharon Adamski, Kurt D’Andrea, Dr. Allison Greenplate and Dr. Amy Baxter are acknowledged for day-to-day running of the CPU, sample processing, and management of the extensive patient sample bank. We thank Lester Lledo and Joan Gilmore in the Clinical Trials Unit of the Center for Cellular Immunotherapies, and Dr. Laurel Glaser for assistance with data compilation, as well as Drs. Sara Cherry and Ronald Collman for helpful advice.

## Notes

### Competing Interest Statement

Joseph A. Fraietta and Carl H. June receive research funding supported in part by a grant from the Parker Institute for Cancer Immunotherapy.

### Clinical Trial

ClinicalTrials.gov: NCT04412785

### Funding Statement

This study was supported through internal funding from the Center for Cellular Immunotherapies at the University of Pennsylvania.

